# Quantifying changes in ventilation for photon- and proton-treated patients according to dose and pneumonitis development

**DOI:** 10.1101/2025.03.06.25323515

**Authors:** Rebecca Lim, Tien Tang, Austin Castelo, Caleb O’Connor, Yulun He, Uwe Titt, Zhongxing Liao, Radhe Mohan, Kristy Brock

## Abstract

Treatment of non-small cell lung cancer with proton therapy allows for delivery of high radiation dose while sparing critical normal tissue structures compared to conventional photon therapy. However, there are currently no established clinical endpoints to study the biological effect between these radiation modalities. Computed tomography (CT) imaging allows for extraction of ventilation through deformable image registration of 4DCTs and may provide information on imaging-based functional changes in the lung. We hypothesize that patients treated with protons exhibit different changes in ventilation compared to those treated with photons. For 20 photon and 20 proton cases, ventilation was extracted using 4DCT Jacobian-based methods and correlated with dose and radiation-induced pneumonitis development. Ventilation was tracked over the course of treatment on a voxel-wise basis and stratified into high and low function. Ventilation differences across modalities were compared by function and pneumonitis development. Patients treated with protons exhibited statistically significant improvements in ventilation change of low functioning voxels compared to photon patients. Future work will evaluate ventilation change specifically in the lung regions that developed pneumonitis.

## 1 Introduction

Lung cancer is the leading cause of cancer death and the second most common form of cancer in the U.S [1]. 80-85% of lung cancer is non-small cell lung cancer (NSCLC) [1], and while surgical resection provides the best outcomes, radiotherapy (RT) is a more feasible option for many patients, especially those advanced in age. However, the lungs are particularly radiosensitive, and RT often leads to toxicities such as radiation-induced pneumonitis (RP) which reduce patient quality of life after treatment [2], [3].

Proton RT is a promising option to reduce normal tissue toxicity, but research to date has failed to demonstrate a clear difference in toxicity development between patients treated with photon or proton RT [4], [5]. Current clinical practice for toxicity modeling and outcome prediction in proton RT relies solely on regional dosimetric factors, such as dose volume histograms (DVHs) and does not incorporate patient-specific biological factors. However, since proton RT is known to produce a different biological effect compared to photon RT [6], [7], [8], biological factors should be considered when designing toxicity models.

Computed tomography ventilation imaging (CTVI) uses deformable image registration (DIR) of 4DCT images to calculate ventilation and has emerged as an efficient alternative to typical ventilation imaging such as single-photon emission computed tomography (SPECT) or positron emission tomography (PET). It has been validated in both animal and human studies [9], [10], and clinical trials have begun to show the potential of using CTVI to design treatment plans that reduce dose to highly functional areas of the lung for toxicity reduction [11], [12]. In particular, it has been shown that pre-treatment ventilation is correlated with dose and pneumonitis development for photon patients [13], [14]. While research has shown that CTVI can be feasibly incorporated into functional avoidance plans for proton therapy [15] and can improve toxicity models for photon patients [16], [17], [18], toxicity models for proton RT have not yet incorporated such functional changes to predict outcomes at a voxel level.

In this study, we extracted ventilation from 4DCTs using Jacobian-based ventilation for both photon- and proton-treated patients. We evaluated how each patient’s ventilation changes on a voxel-wise basis over the course of treatment across treatment modality, RP development, and dose. We hypothesize that there will be significant differences in ventilation change over treatment across radiation modalities. In particular, since proton RT is known to reduce normal tissue toxicity, we hypothesize that proton-treated patients will have a greater capacity for ventilation improvement due to greater sparing of normal lung tissue.

## 2 Materials and Methods

40 patients treated for NSCLC were chosen retrospectively from a randomized clinical trial [4]. 20 patients were treated with photon RT, 12 of which developed RP, and 20 patients were treated with proton RT, 10 of which developed RP. Photon patients were treated with intensity modulated radiation therapy (IMRT) and proton patients were treated with passive scattering proton therapy (PSPT), each for 37 fractions at a prescription dose of 74 Gy. All patients received concurrent chemotherapy. Each patient had 4DCTs acquired for planning and weekly up to the final week of treatment, as well as a PET/CT scan acquired 3-12 months after treatment, which was used to diagnose RP. Dose plans for each patient were calculated on the exhale (T5) scan of the planning week.

The Jacobian determinant, shown below, is a representation of local volume change and can be calculated using DIR [9].

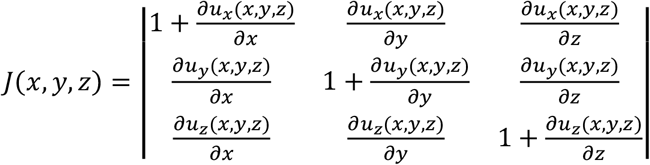

The Jacobian determinant of the displacement vector field (DVF) from each registration was used to determine each voxel’s differential expansion or contraction, enabling the calculation of the specific volume change of the voxel, *J(x,y,z) – 1*. An additional scaling factor using the Hounsfield Unit (HU) values of the associated T5 scan of the 4DCT was incorporated to provide a more physiologically accurate measurement of ventilation and converted specific volume change to specific ventilation [19], denoted by *V*:

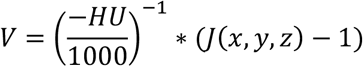

Figure 1 shows the workflow for determining voxel-wise correlation of the Jacobian over treatment. First, intra-fraction DIR using a hybrid intensity-based algorithm was performed between the T0 and T5 scans of the 4DCTs for the planning week and final week of treatment separately. Focus ROIs were used to align the lung contours during the registration process. The resulting intra-fraction DIRs were used to calculate the Jacobian determinant. Next, an inter-fraction DIR was performed between the T5 scans of the planning and final weeks of treatment. This step allowed for voxel-wise correspondence across timepoints. For photon patients, a biomechanical-based algorithm was used to perform the inter-fraction DIR, and a hybrid intensity-based algorithm was used for the proton patients. The DVF from the inter-fraction DIR was applied to the Jacobian map from the final week to map it to the planning week coordinate space. The resulting Jacobian maps were then scaled using the HU correction factor to calculate ventilation. Since the planned doses were also done on the T5 scan of the planning week, the dose maps match the Jacobian maps on a voxel level. Lastly, the gross tumor volume (GTV) was segmented using an in-house segmentation model and removed from the evaluation to avoid erroneous ventilation calculations. Grills et al. found that GTV contours underestimate composite pathologic tumor size by 1.2 mm [20], so all GTV contours were expanded by 3 mm to ensure consistency before removal from the lung contour.

**Figure 1:**
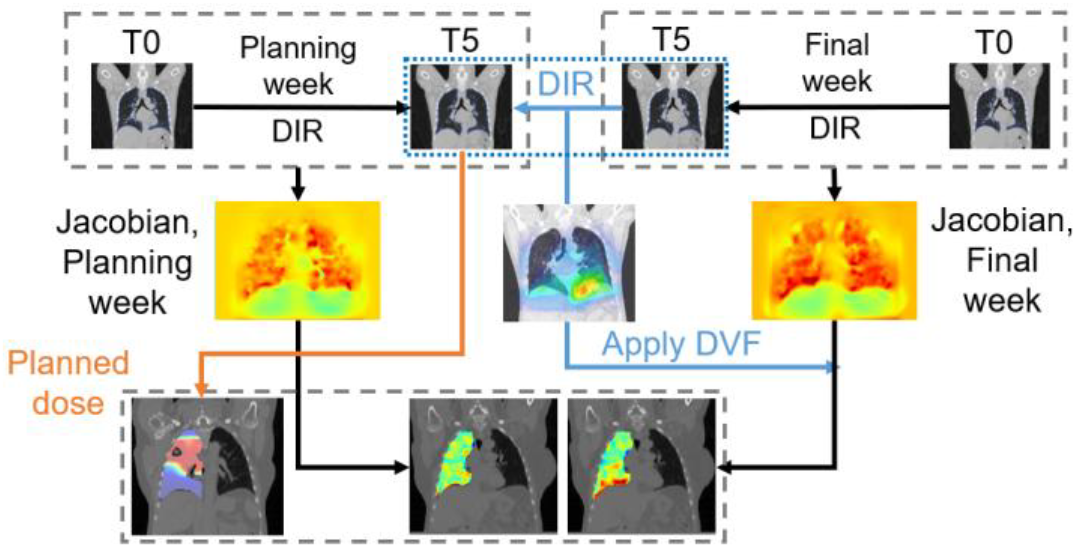
Workflow figure to extract voxel-wise change in ventilation over the course of treatment and accumulated dose using deformable image registration (DIR) and deformation vector fields (DVFs).

The change in ventilation between beginning and end of treatment for each voxel was calculated for each patient. Voxels were then stratified by function, toxicity, and treatment modality. To stratify voxels by function, the median Jacobian value was determined from the planning week ventilation map for each patient. A high functioning voxel was defined as a voxel that had a ventilation value above the median value in the planning week, and a low functioning voxel was defined to have a ventilation value below the median value in the planning week. After stratifying by function, voxels were binned by dose in 5 Gy increments and the ventilation differences were averaged in each bin. The averaged differences were plotted against dose for proton and photon patients, split into RP vs. non-RP patients for all voxels, then split again into high vs. low functioning voxels. T-tests were conducted on the ventilation differences between photon and proton patients, within each dose bin as well as across all dose bins.

## 3 Results

Boxplots of the average differences in ventilation for photon and proton patients are shown in Figure 2. Average differences further split into high and low functioning voxels are shown in Figure 3.

**Figure 2:**
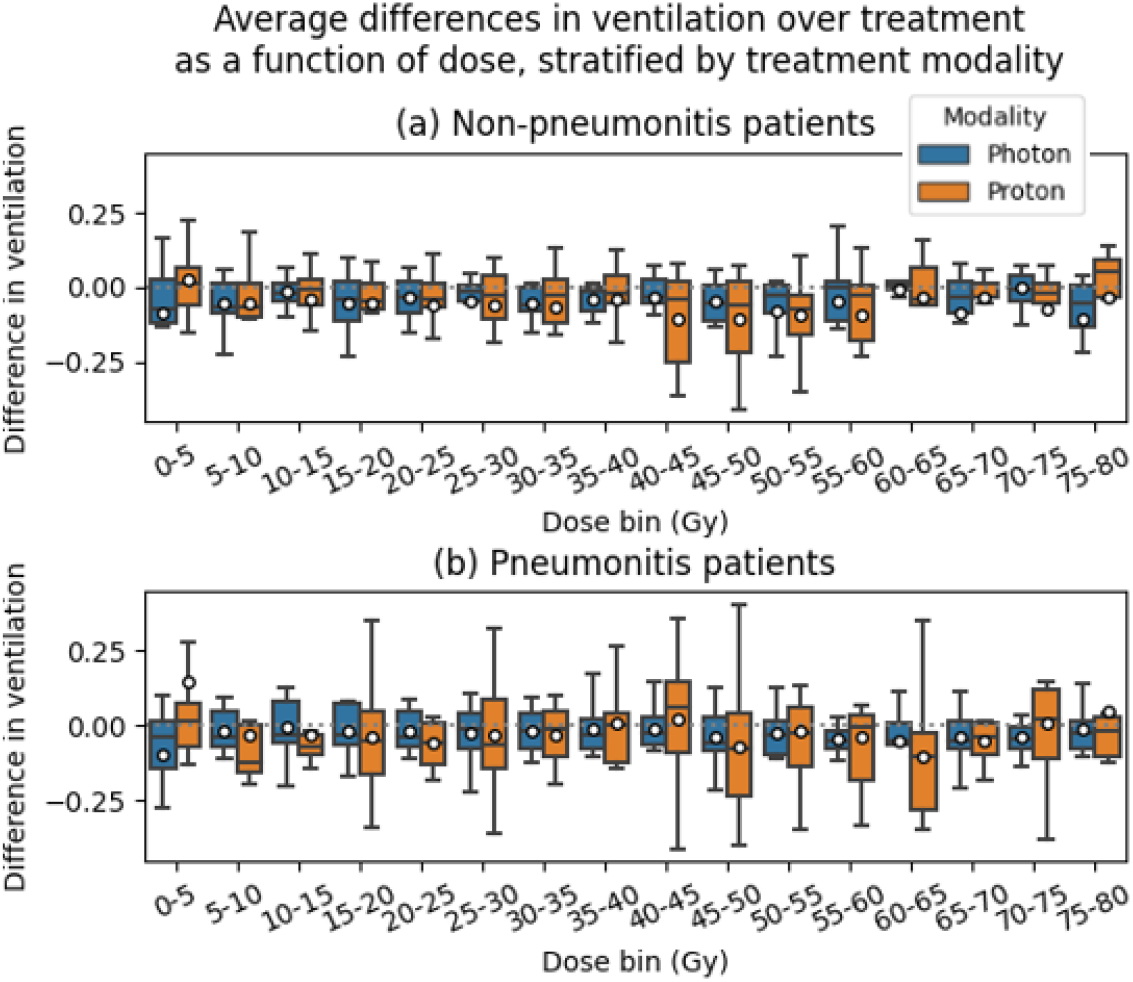
Boxplots of average differences in ventilation for patients that did not develop pneumonitis (a) and patients that did (b). Patients were split into photon and proton cohorts, shown in blue and orange respectively. Dose bins are 5 Gy in width and range from 0 to 80 Gy. Ventilation function is determined by the median of the ventilation values from the planning week.

**Figure 3:**
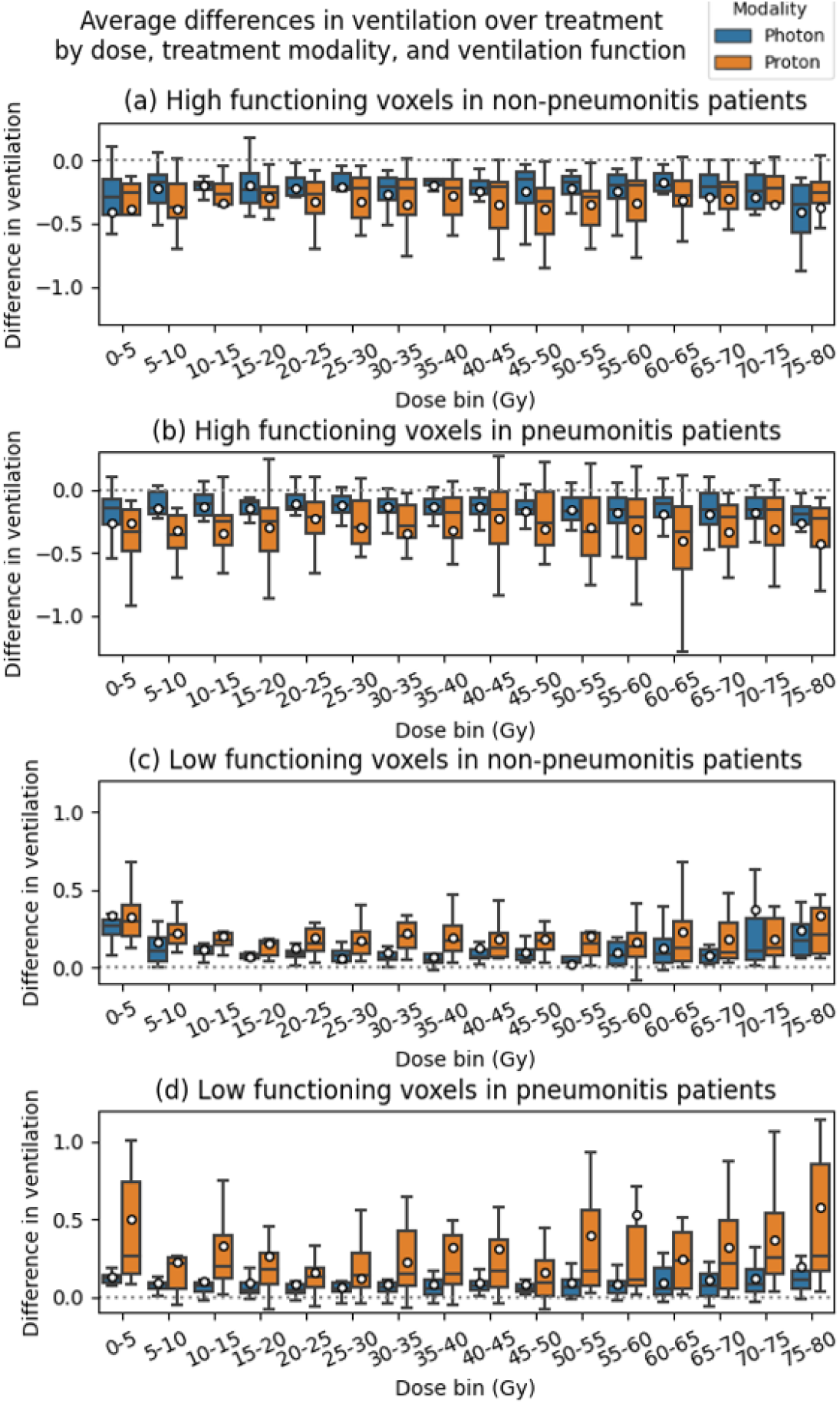
Boxplots of average differences in ventilation for pneumonitis (RP) and non-RP patients, split by toxicity and ventilation function. (a) and (b) show high functioning voxels in patients that did not and did develop RP respectively. (c) and (d) show low functioning voxels in patients that did not and did develop RP. Patients were split into photon and proton cohorts, shown in blue and orange respectively. Dose bins are 5 Gy in width and range from 0 to 80 Gy. Ventilation function is determined by the median of the ventilation values from the planning week.

High functioning voxels decreased in ventilation over the course of treatment while low functioning voxels increased in ventilation, with relatively small changes in overall ventilation averaged across function. In the high functioning voxels, ventilation tended to decrease by a greater magnitude for proton treated patients compared to photon treated patients. In contrast, proton patients demonstrated an increase in ventilation in low functioning voxels where photon treated patients did not.

Ventilation differences for all voxels prior to stratification by function were not found to be significant. T-test p-values for the high and low functioning voxels corresponding to Figure 2 are shown in Table 1. Ventilation differences were found to be statistically significant (p < 0.05) when considering all dose bins. Within each dose bin, statistical significance was found only in the low functioning voxels. In particular, the 15-20 Gy, 25-30 Gy, and 30-35 Gy dose bins were found to be statistically significant when comparing patients that did not develop RP, and the 70-75 Gy dose bin was significant when comparing patients that developed RP.

**Table 1:** P-values for t-tests conducted across treatment modalities. T-tests were performed for each cohort within each dose bin and over all dose bins combined. Statistically significant p-values are shown in bold.

| Dose bin (Gy) | Low function, non-RP, photon vs. proton | Low function, RP, photon vs. proton | High function, non-RP, photon vs. proton | High function, RP, photon vs. proton |
| --- | --- | --- | --- | --- |
| 0-5 | 0.923 | 0.061 | 0.9097 | 0.989 |
| 5-10 | 0.408 | 0.137 | 0.1582 | 0.247 |
| 10-15 | 0.063 | 0.062 | 0.1107 | 0.064 |
| 15-20 | **0.032** | 0.161 | 0.3099 | 0.156 |
| 20-25 | 0.299 | 0.309 | 0.2504 | 0.149 |
| 25-30 | **0.030** | 0.667 | 0.2977 | 0.105 |
| 30-35 | **0.037** | 0.111 | 0.5015 | 0.093 |
| 35-40 | **0.019** | 0.172 | 0.3334 | 0.375 |
| 40-45 | 0.413 | 0.160 | 0.3549 | 0.419 |
| 45-50 | 0.327 | 0.319 | 0.2546 | 0.377 |
| 50-55 | 0.058 | 0.067 | 0.1319 | 0.209 |
| 55-60 | 0.430 | 0.200 | 0.4536 | 0.310 |
| 60-65 | 0.304 | 0.054 | 0.1033 | 0.172 |
| 65-70 | 0.090 | 0.088 | 0.9728 | 0.285 |
| 70-75 | 0.415 | **0.047** | 0.6856 | 0.313 |
| 75-80 | 0.551 | 0.100 | 0.8147 | 0.376 |
| All bins | **0.003** | **1.76E-08** | **0.003** | **1.96E-05** |

## 4 Discussion

Statistical significance was found when comparing all dose bins, indicating that there is a significant difference in ventilation overall when comparing modalities. High functioning voxels exhibited a decrease in ventilation over the course of treatment for all patients while low functioning voxels increased over the course of treatment. These findings were similarly demonstrated in Patton et al. [13], who showed that higher functioning voxels decreased by 4.8% per 10 Gy compared to lower functioning voxels, which decreased by 2.4%. Additionally, statistical significance within dose bins was found mostly above 20 Gy, which has also been shown to be a significant threshold for toxicity development by King et al. [21] and Graham et al. [22].

Proton patients tended to have more extreme changes in ventilation overall as demonstrated by the t-tests. In the low functioning voxels, the increase in ventilation was significantly higher for proton patients, supporting the hypothesis that the higher sparing of normal tissue in proton RT allows for greater increase in function.

Some limitations of this study include the lack of spatial information for each voxel and the large amounts of averaging for each patient. Future work will include a more in-depth analysis of the voxels within the regions of the lung that developed RP for patients that were diagnosed with RP. Since different DIR strategies were employed across time points for photon patients compared to proton patients, we will also be comparing results achieved with the same interfraction DIR. The photon patients in our dataset also have accumulated dose plans, and a future study will compare how ventilation changes for planned and accumulated doses. Lastly, additional patients will be incorporated for further analysis.

## 5 Conclusion

We developed a workflow to calculate voxel-wise change in lung ventilation and dose using Jacobian-based methods. Proton patients exhibited more extreme changes in ventilation in comparison to photon patients. Future work will incorporate more patients and work towards establishing CT based lung ventilation as a functional endpoint when evaluating differences between photon and proton therapy.

## Data Availability

All data produced in the present study are available upon reasonable request to the authors in compliance with institutional IRB requirements.

## Acknowledgements

Research reported in this publication was supported in part by the National Cancer Institute of the National Institutes of Health under award numbers P30CA016672 and P01CA261669. Research reported in this publication was supported in part by resources of the Image Guided Cancer Therapy Research Program at The University of Texas MD Anderson Cancer Center.

## References

[1] R. L. Siegel, K. D. Miller, N. S. Wagle, and A. Jemal, “Cancer statistics, 2023,” CA A Cancer J Clinicians, vol. 73, no. 1, pp. 17– 48, Jan. 2023, doi: 10.3322/caac.21763.

[2] X.-J. Zhang et al., “Prediction of radiation pneumonitis in lung cancer patients: a systematic review,” J Cancer Res Clin Oncol, vol. 138, no. 12, pp. 2103–2116, Dec. 2012, doi: 10.1007/s00432-012-1284-1.

[3] M. Aiad et al., “Comparison of Pneumonitis Rates and Severity in Patients With Lung Cancer Treated by Immunotherapy, Radiotherapy, and Immunoradiotherapy,” Cureus, Jun. 2022, doi: 10.7759/cureus.25665.

[4] Z. Liao et al., “Bayesian Adaptive Randomization Trial of Passive Scattering Proton Therapy and Intensity-Modulated Photon Radiotherapy for Locally Advanced Non–Small-Cell Lung Cancer,” JCO, vol. 36, no. 18, pp. 1813–1822, Jun. 2018, doi: 10.1200/JCO.2017.74.0720.

[5] S. Mesko and D. Gomez, “Proton Therapy in Non-small Cell Lung Cancer,” Curr. Treat. Options in Oncol., vol. 19, no. 12, p. 76, Nov. 2018, doi: 10.1007/s11864-018-0588-z.

[6] H. Paganetti, “Relative biological effectiveness (RBE) values for proton beam therapy. Variations as a function of biological endpoint, dose, and linear energy transfer,” Phys. Med. Biol., vol. 59, no. 22, p. R419, Oct. 2014, doi: 10.1088/0031-9155/59/22/R419.

[7] E. T. Vitti and J. L. Parsons, “The Radiobiological Effects of Proton Beam Therapy: Impact on DNA Damage and Repair,” Cancers, vol. 11, no. 7, Art. no. 7, Jul. 2019, doi: 10.3390/cancers11070946.

[8] R. Mohan and D. Grosshans, “Proton therapy – Present and future,” Advanced Drug Delivery Reviews, vol. 109, pp. 26–44, Jan. 2017, doi: 10.1016/j.addr.2016.11.006.

[9] J. M. Reinhardt, K. Ding, K. Cao, G. E. Christensen, E. A. Hoffman, and S. V. Bodas, “Registration-based estimates of local lung tissue expansion compared to xenon CT measures of specific ventilation,” Medical Image Analysis, vol. 12, no. 6, pp. 752–763, Dec. 2008, doi: 10.1016/j.media.2008.03.007.

[10] “The VAMPIRE challenge: A multi‐institutional validation study of CT ventilation imaging - Kipritidis - 2019 - Medical Physics - Wiley Online Library.” Accessed: Feb. 02, 2024. [Online]. Available: https://aapm.onlinelibrary.wiley.com/doi/full/10.1002/mp.13346

[11] E. Vicente et al., “Functionally weighted airway sparing (FWAS): a functional avoidance method for preserving post-treatment ventilation in lung radiotherapy,” Phys. Med. Biol., vol. 65, no. 16, p. 165010, Aug. 2020, doi: 10.1088/1361-6560/ab9f5d.

[12] T. Yamamoto et al., “Four-Dimensional Computed Tomography Ventilation Image-Guided Lung Functional Avoidance Radiation Therapy: A Single-Arm Prospective Pilot Clinical Trial,” International Journal of Radiation Oncology*Biology*Physics, vol. 115, no. 5, pp. 1144–1154, Apr. 2023, doi: 10.1016/j.ijrobp.2022.11.026.

[13] T. J. Patton, S. E. Gerard, W. Shao, G. E. Christensen, J. M. Reinhardt, and J. E. Bayouth, “Quantifying ventilation change due to radiation therapy using 4 DCT Jacobian calculations,” Medical Physics, vol. 45, no. 10, pp. 4483–4492, Oct. 2018, doi: 10.1002/mp.13105.

[14] M. Otsuka, H. Monzen, N. Kadoya, M. Inada, K. Matsumoto, and Y. Nishimura, “Evaluation of Lung Toxicity Risk with Computed Tomography Ventilation Functional Image for Lung Stereotactic Body Radiation Therapy and Three-Dimensional Conformal Radiation Therapy,” International Journal of Radiation Oncology*Biology*Physics, vol. 99, no. 2, Supplement, p. E708, Oct. 2017, doi: 10.1016/j.ijrobp.2017.06.2306.

[15] Q. Huang et al., “Dosimetric feasibility of 4DCT-ventilation imaging guided proton therapy for locally advanced non-small-cell lung cancer,” Radiat Oncol, vol. 13, no. 1, p. 78, Apr. 2018, doi: 10.1186/s13014-018-1018-x.

[16] A. M. Faught et al., “Evaluating Which Dose-Function Metrics Are Most Critical for Functional-Guided Radiation Therapy,” International Journal of Radiation Oncology*Biology*Physics, vol. 99, no. 1, pp. 202–209, Sep. 2017, doi: 10.1016/j.ijrobp.2017.03.051.

[17] Y. Vinogradskiy et al., “Use of 4-Dimensional Computed Tomography-Based Ventilation Imaging to Correlate Lung Dose and Function With Clinical Outcomes,” International Journal of Radiation Oncology*Biology*Physics, vol. 86, no. 2, pp. 366–371, Jun. 2013, doi: 10.1016/j.ijrobp.2013.01.004.

[18] A. M. Faught et al., “Evaluating the Toxicity Reduction With Computed Tomographic Ventilation Functional Avoidance Radiation Therapy,” International Journal of Radiation Oncology*Biology*Physics, vol. 99, no. 2, pp. 325–333, Oct. 2017, doi: 10.1016/j.ijrobp.2017.04.024.

[19] R. Castillo, E. Castillo, J. Martinez, and T. Guerrero, “Ventilation from four-dimensional computed tomography: density versus Jacobian methods,” Phys. Med. Biol., vol. 55, no. 16, pp. 4661– 4685, Aug. 2010, doi: 10.1088/0031-9155/55/16/004.

[20] I. S. Grills et al., “Clinicopathologic Analysis of Microscopic Extension in Lung Adenocarcinoma: Defining Clinical Target Volume for Radiotherapy,” International Journal of Radiation Oncology*Biology*Physics, vol. 69, no. 2, pp. 334–341, Oct. 2007, doi: 10.1016/j.ijrobp.2007.03.023.

[21] M. T. King, P. G. Maxim, M. Diehn, B. W. Loo, and L. Xing, “Analysis of Long-Term 4-Dimensional Computed Tomography Regional Ventilation After Radiation Therapy,” Int J Radiat Oncol Biol Phys, vol. 92, no. 3, pp. 683–690, Jul. 2015, doi: 10.1016/j.ijrobp.2015.02.037.

[22] M. V. Graham et al., “Clinical dose-volume histogram analysis for pneumonitis after 3D treatment for non-small cell lung cancer (NSCLC),” Int J Radiat Oncol Biol Phys, vol. 45, no. 2, pp. 323– 329, Sep. 1999, doi: 10.1016/s0360-3016(99)00183-2.

